# Linear ablation for persistent atrial fibrillation: an evidence-based study

**DOI:** 10.1101/2023.08.30.23294868

**Authors:** Changhao Xu, Kaige Li, Xiyao Zhu, Xinhua Wang, Ping Ye, Weifeng Jiang, Shaohui Wu, Kai Xu, Xiangting Li, Ying Wang, Qidong Zheng, Yanzhe Wang, Lihua Leng, Zengtang Zhang, Bing Han, Yu Zhang, Mu Qin, Xu Liu

**Affiliations:** Department of Cardiology, Shanghai Chest Hospital, Shanghai Jiao Tong University, Shanghai, China; Department of Clinical Integration of Traditional Chinese and Western medicine, First Clinical Medical College, Shandong University of Traditional Chinese Medicine, No.4655 University Road, Changqing District, Jinan City, Shandong 250355, China; Department of Cardiology, Ren Ji Hospital, Shanghai Jiao Tong University School of Medicine, No.160 Pujian Road, Shanghai 200127, China; Department of Cardiology, The Central Hospital of Wuhan, Tongji Medical College, Huazhong University of Science and Technology, No.26 Shengli Street, Jiang’an District, Wuhan City, Hubei 430014, China; Department of Cardiology, Affiliated Hospital of Jining Medical University, No.89 Guhuai Road, Jining City, Shandong 272007, China; Department of Cardiology, Second Affiliated Hospital of Shandong University of Traditional Chinese Medicine, No.1 Jingba Road, Jinan City, Shandong 25000, China; Department of Cardiology, Yuhuan Second People’s Hospital, No.77 Huanbao Road, Yuhuan City, Zhejiang 317600, China; Department of Cardiology, Changshu Hospital of Traditional Chinese Medicine, No. 6 Huanghe Road, Changshu City, Jiangsu 215516, China; Department of Cardiology, The PLA Navy Anqing Hospital, No.150 Shuangjing Street, Anqing City, Anhui 246003, China; Department of Cardiology, Jinan City People’s Hospital, No.1 Xuehu Street, Changshao North Road, Laiwu District, Jinan City, Shandong 250102, China; Department of Cardiology, Xuzhou Central Hospital, No.199 South Jiefang Road, Xuzhou City, Jiangsu 221009, China

**Keywords:** Key word: Atrial fibrillation, Catheter ablation, Atrial tachycardia, Linear ablation

## Abstract

**Background:** Additional linear ablation for persistent atrial fibrillation (PerAF) still has limited evidence-based medical proof.

**Objectives:** We probed into the mechanisms of intermediate atrial tachycardia (AT) during PerAF termination by catheter ablation and provided evidence for it.

**Methods:** 136 patients who converted to organized AT after PerAF termination in the Extent-AF study were analyzed. Bi-atrial activation mapping combined with entrainment mapping were performed to identify the mechanisms and critical isthmus of these ATs.

**Results:** A total of 164 ATs in 136 patients were identified (average 1.2 per patients) and 143 (87%) ATs in 113 patients (average 1.3 per patient) were successfully mapped. The mechanisms of intermediate ATs were macro-reentry in 110 (77%), micro-reentry in 21 (15%), and focal AT in 12 (8%). Among the macro-reentrant ATs, the most common were perimitral ATs (PM-AT) 52 (47%), followed by roof dependent ATs (RF-AT) in 40 (36%) and typical atrial flutter (AFL) in 18 (16%). 98 (72%) patients had successfully ablated intermediate ATs. Among these patients, 88 (90%) required at least one of the perimitral line, roofline, or peritricuspid line to finally restore sinus rhythm. At the end of 12 months of follow-up, 63 (64.3%) patients with successful ablative ATs were free of any arrhythmia.

**Conclusion:** The majority of intermediate ATs after PerAF termination were macro-reentrant ATs. Linear ablation targets the mitral isthmus, roof, and tricuspid isthmus was a critical step of PerAF ablation to restore sinus rhythm in up to 90% patients, suggesting the importance of additional linear ablation.

## Introduction

Despite optimal strategy for persistent atrial fibrillation (PerAF) ablation have not been well established, linear ablation was still the most popular additional ablative strategy since 1994(1), which has substrate compartmentalization effect that reduces the area available for the completion of wavelet circuits. Among the several linear lesions, the perimitral line and the roofline were most widely used. The rationale for mitral isthmus ablation is the preferential propagation circumferential around the mitral annulus that may facilitate AF in response to functional or anatomic conduction block(2). In addition, left atrial roof is a region with highly fragmented electrograms and plays a role in supporting macro-reentry around the pulmonary veins, which is the theoretical basis for roof line ablation(3). Despite several theories support the rationality of performing additional linear ablation, it still lacks evidence-based medical proof and its effectiveness remains to be verified. The Extent-AF study demonstrated that procedural termination of PerAF indicates a higher probability of sinus rhythm maintanence(4). Study of us and other groups showed that up to 87% PerAF converted to organized atrial tachycardia (AT) before sinus rhythm (SR) restoration during catheter ablation(4–7). Furthermore, successful ablation of intermediate ATs led to better rhythm outcome in PerAF patients(8). Therefore, the intermediate ATs, a transient state between PerAF and SR, may represent remnant arrhythmic mechanism after AF termination. It is plausible that the mechanisms of intermediate ATs may provide evidence-based medical proof for appropriate linear ablation. The Extent-AF study was a large prospective, multicenter trial comparing three mainstream ablation strategies in patients with PerAF(4). We aim to investigate the clinical significance of ATs after PerAF termination in the Extent-AF study.

## Methods

### Study population

We enrolled 136 patients who converted to organized AT after PerAF termination in the Extent-AF study. The detailed study protocol has been published previously(4). The study was approved by the ethics committees at each participant center.

### Study procedures

All patients were in spontaneous AF at the beginning of the procedure. Patients were randomized assigned to receive the following three ablation strategies in a 1:1:1 ratio, which included: PVI plus linear ablation and/or vein of Marshall (VOM) ethanol infusion, PVI plus electrogram (EGM)-guided ablation, extensive ablation (PVI plus anatomical-guided ablation and EGM-guided ablation) PVI was successfully performed in all patients. Catheter ablation was performed under the guidance of electro-anatomical mapping system (CARTO, Biosense Webster, CA, USA). Targeted EGM includes locally short-cycle length potentials, spatial-temporal dispersion potentials, high-frequency potentials, and focal activity.

If AF converted to stable AFL/AT, atrial activation mapping combined with multiple sites entrainment mapping were performed to identify the critical isthmus of these ATs. A high-density mapping technique was employed in all cases. To annotate electrograms during activation mapping, a stable intracardiac reference is necessary. Typically, a multipolar catheter placed in the coronary sinus (CS) serves this purpose. However, if there is instability or the presence of low-voltage atrial or large far-field ventricular electrograms, the reference catheter can be placed in one of the appendages. The “window-of-interest (WOI)” was set to encompass approximately 90% of the cycle length (CL) to capture tachycardias with various mechanisms. Mapping of the left atrium was performed initially using a Pentaray catheter. The mapping results were interpreted as follows:

1. The presence of common macro-reentry patterns, such as mitral annular flutter, roof-dependent flutter, or scar-related reentry, was assessed. This involved determining whether there was a conspicuous “red-to-purple” transition in the corresponding anatomical regions (around the mitral annulus, roof, or near scar tissue). If this transition was observed, the atrial flutter was initially classified as macro-reentry. Subsequently, entrainment mapping was performed at relevant key anatomical sites, including the anterior wall, left atrial roof, left atrial bottom, left atrial appendage base, mitral isthmus, left atrial posterior wall, and areas adjacent to scar tissue, to validate the findings. Entrainment mapping was conducted by applying pacing stimuli with a cycle length 20 to 30ms shorter than the tachycardia cycle length (TCL). A site was considered part of the reentry circuit if the post-pacing interval (PPI), measured from the stimulation artifact to the return atrial electrogram on the ablation catheter, fell within 30ms of the TCL.
2. The possibility of wavefront collision around the mitral valve, roof, or tricuspid valve, which would exclude perimitral atrial flutter, roof-dependent atrial flutter, or tricuspid valve-dependent atrial flutter, respectively, was evaluated. If these conditions were ruled out, special types of atrial flutter, such as localized reentry or focal activation, were considered. Careful mapping of specific atrial electrograms, including areas with long-duration signals and fragmented potentials, was performed. The electrogram activation sequence was adjusted based on the operator’s experience. Focal AT was maintained by activity originating from a localized region with a centrifugal activation pattern recorded in the electro-anatomical mapping system. Additionally, voltage mapping information was used to identify suspicious areas of low voltage. It is worth noting that regions exhibiting complex electrograms often correspond to critical isthmuses for these types of atrial flutter. Entrainment mapping techniques (as described earlier) were applied in other regions of the left atrium to further exclude the possibility of macro-reentry and determine the spatial distribution of the left atrial reentry circuit.
3. If unsatisfactory results were obtained from entrainment mapping within the left atrium or if there were time gaps exceeding 20ms, passive activation within the left atrium was considered, suggesting the potential presence of the main reentry circuit in the right atrium. In such cases, activation mapping and entrainment mapping were performed in the right atrium, following the same procedures as in the left atrium. Clear mapping results and effective entrainment pacing in the right atrium would confirm the location of the main reentry circuit in the right atrium. If time gaps in activation were also observed in the right atrium, the possibility of bi-atrial reentry or extracardiac involvement in atrial flutter should be considered.

For macro reentrant ATs, the key isthmus was ablated and conduction block of the linear lesions were confirmed using pacing protocol after sinus rhythm (SR) restoration. If perimetral block cannot be achieved by endocardial ablation, ablation inside the coronary sinus or VOM ethanol infusion was performed. Similarly, key isthmus was also ablated in localized ATs. If the patient has multiple mechanisms of ATs, the operator should pursue successful ablation of each type of AT.

The procedure should be stopped and 150-200 J direct current conversion (DCCV) was performed to restore SR under the following situation: 1) for safety reasons; 2) AF still persists despite all ablation lesions were created appropriately; 3) the procedural time was beyond 5 hours; 4) difficulties associated with successfully identify the mechanisms of the ATs, eliminating all the marked EGM area, or creating a complete bidirectional block linear lesion despite the operator’s best efforts. Bidirectional block of the linear lesions was verified in SR.

### Follow-up

Anti-arrhythmic medications were discontinued in all patients before 3 months. The first 3 months after the initial ablation was considered as a “blanking period”. Outpatient visits and 48h Holter monitoring were scheduled at 1, 3, 6, 9, 12 months, and every 6 months thereafter if the patient remained asymptomatic. Monthly telephone interviews were also done. All patients were strongly recommended to undergo additional ECGs and 7-day Holter recordings when their symptoms were suggestive of tachycardia. A “recurrence” of atrial arrhythmia was considered any episode lasting 30s (symptomatic or asymptomatic) detected by ECG and/or Holter.

### Statistical analysis

Continuous variables were expressed as mean ± standard deviation or median with interquartile range and compared using independent sample t-tests or non-parametric tests, whereas discrete variables were expressed as percentages and compared using x2 tests.

Survival curves were performed using the Kaplan–Meier method.All tests of significance were two-sided, with a probability <0.05 considered significant. All statistical analyses were performed using SPSS version 9.1 (SAS institute, Inc., Cary, NC) and GraphPad Prism 9.0 software (GraphPad Software Inc, San Diego, CA, USA).

## Results

### Study population

The extent-AF study included a total of 450 patients in 10 experienced centers in China. 216 (48%) patients experienced acute AF termination and 63% (136/216) converted to one or more organized AFL/AT. The baseline characteristics of these patients were listed in the **Table 1**.

**Table 1.**
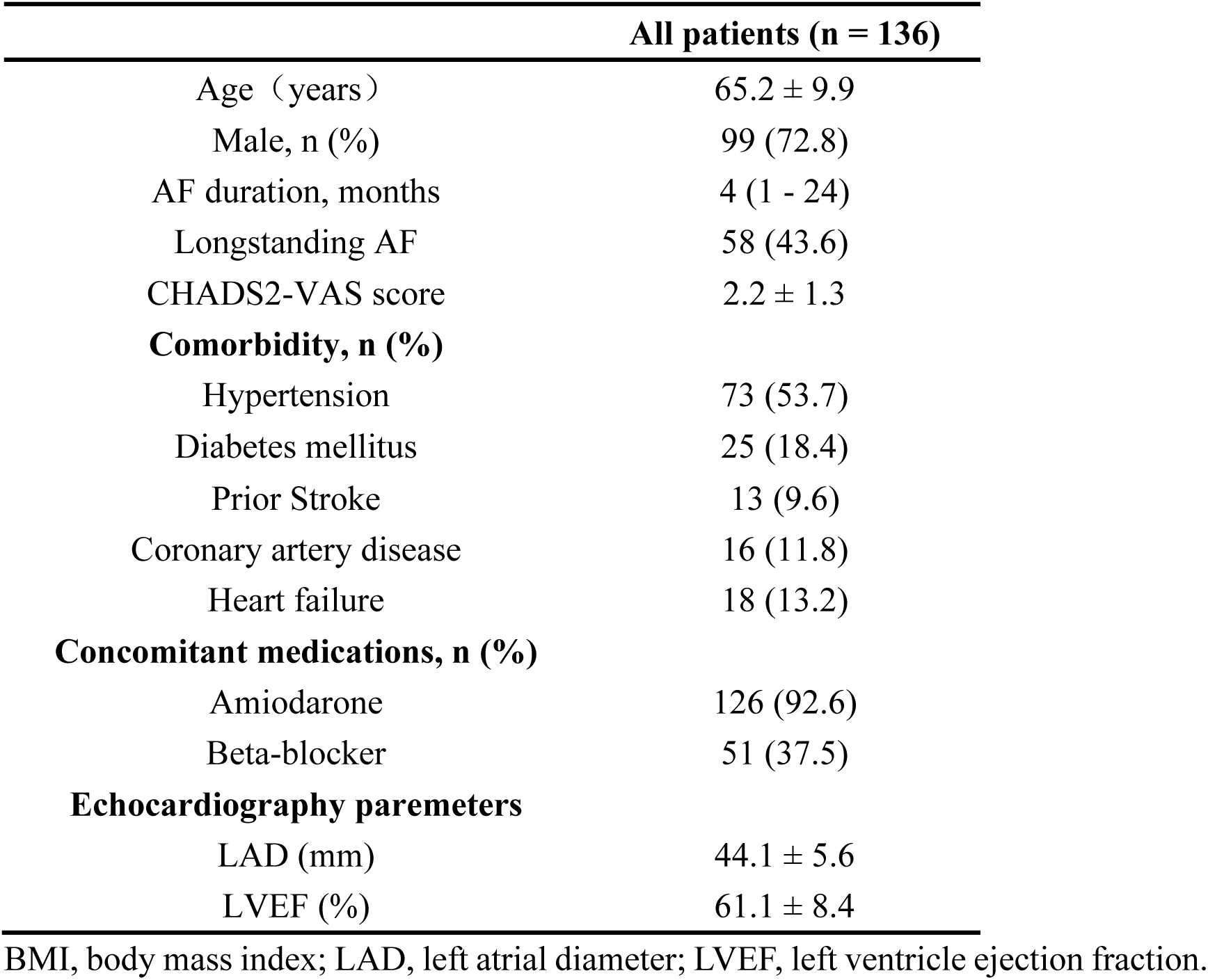
Baseline characteristics.

### Procedural characteristics

PVI was successfully performed in all patients. A total of 164 ATs in 136 patients were identified (average 1.2 per patients). In detail, 11 (7%) types ATs stopped spontaneously and could not be re-induced, 4 (2%) ATs terminated during entrainment mapping, and 8 (5%) had constantly changing TCL during mapping and were unable to identify the mechanism and critical isthmus of the ATs. Therefore, a total of 143 (87%) ATs were successfully mapped in the 113 (average 1.3 per patient) patients.

In the 113 patients with clear mechanisms of ATs, 86 (76%) were presented as single mechanism AT and 27 (24%) had at least two pattens of ATs. 128 (89%) ATs in 98 (87%) patients were successfully ablated. The remanent 15 (13%) patients underwent DCCV to restore SR. The mapping time and radiofrequency (RF) time for single mechanism AT were 19.5 (IQ range 16 – 26.5) min and 15 (IQ range 10 – 20) min, respectively. As for patients who had at least two pattens of ATs, the mapping time and radiofrequency (RF) time were 45 (IQ range 33 – 55) min and 27 (IQ range 22 – 29) min, respectively.

### Mechanism of intermediate ATs

As shown in Table 2. the mechanisms of intermediate ATs were macro-reentry in 110 (77%), micro-reentry in 21 (15%), and focal AT in 12 (8%). Among the macro-reentrant ATs, the most common were perimitral ATs (PM-AT) 52 (47%), followed by roof dependent ATs (RF-AT) in 40 (36%) and typical atrial flutter (AFL) in 18 (16%) (Figure 1B). As for micro-reentrant ATs, the critical sites were left atrial anterior wall in 11 (52%) patients, PV in in 6 (29%), LA appendage ridge in 2 (10%), septum in 2(10%), and superior vena cava in 1 (5%) (Figure 1C). 12 ATs were considered as focal ATs, 7 (58%) were related to the left atrial anterior wall, 3 (25%) were related to PV, and 2 (16%) involved LAA (Figure 1D).

**Figure 1.**
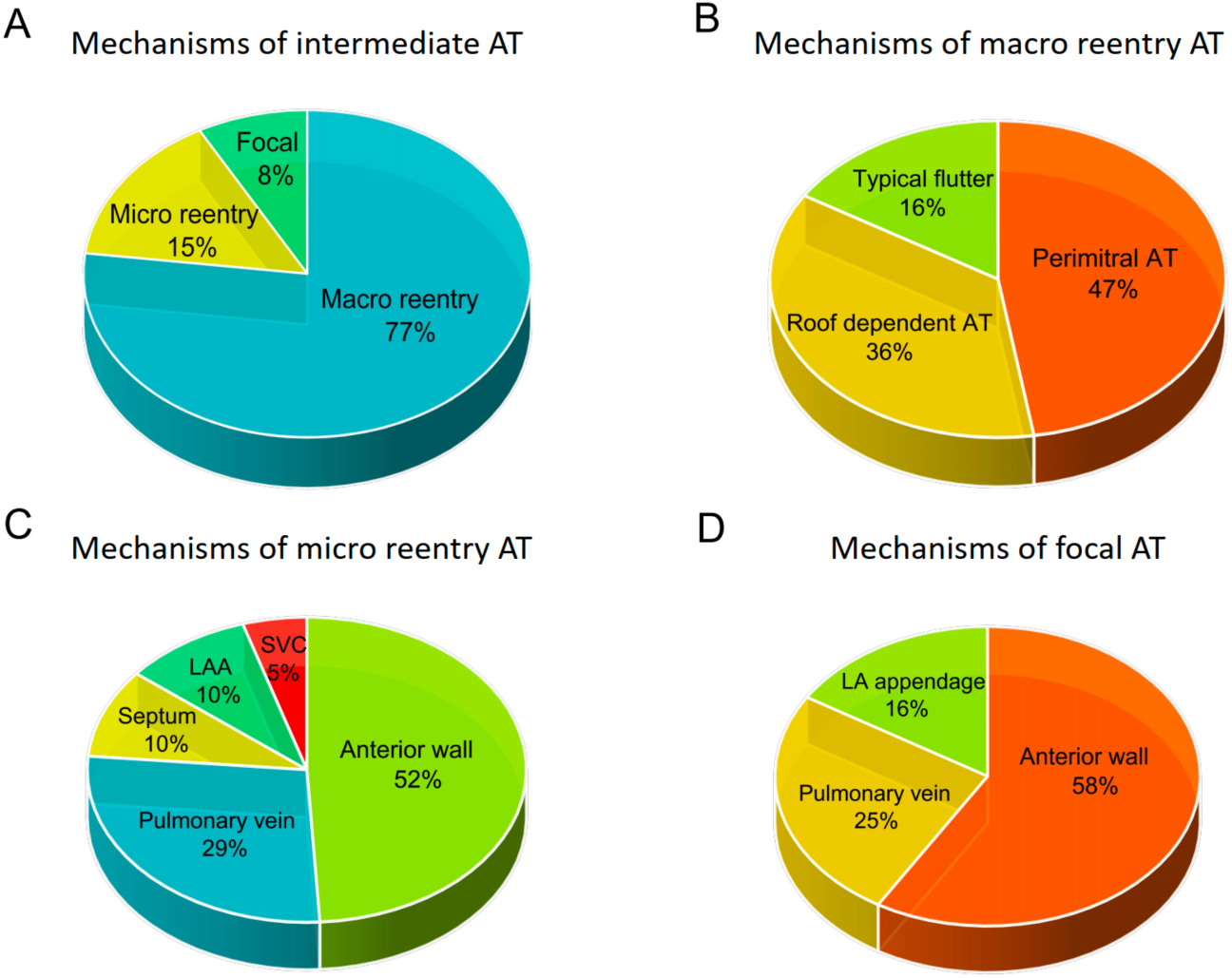
Mechanism of intermediate ATs AT, Atrial Tachycardia; LAA, Left Atrial Appendage; SVC, Superior Vena Cava; LA, Left Atrium.

**Table 2.** Distribution and mechanisms of intermediate ATs.

| AT mechanisms | Critical sites | n (%) |
| --- | --- | --- |
| Macro-reentry |  | 110 (77%) |
|  | Mitral isthmus | 52 (36%) |
|  | Roof | 40 (28%) |
|  | Tricuspid isthmus | 18 (13%) |
| Micro-reentry |  | 21 (15%) |
|  | LA anterior wall | 11 (8%) |
|  | PV | 6 (4%) |
|  | LA appendage | 2 (1%) |
|  | Septum | 2 (1%) |
|  | SVC | 1 (1%) |
| Focal |  | 12 (8%) |
|  | LA anterior wall | 7 (5%) |
|  | PV | 3 (2%) |
|  | LA appendage | 2 (1%) |

6 LA, left atrial; PV, pulmonary vein; SVC, superior vena cava.

### Ablation of single mechanism intermediate ATs

In 86 patients with single mechanism AT, 68 (79%) were presented as macro reentry AT, which included 35 (41%) PM-AT, 23 (27%) RF-AT, and 10 (12%) typical AFL. Endocardial mitral linear ablation only successfully terminated 22 (63%) perimitral ATs. Rate of endocardial perimitral block was 67% by endocardial mitral linear ablation only with an RF application time of 20 min (IQ range 18 – 23). 28% patients underwent epicardial ablation inside the coronary sinus and 40% received VOM ethanol infusion. After epicardial ablation and/or VOM ethanol infusion, the rate of successful primitral block and termination of perimetral ATs increased to 83% and 91%, respectively. Complete block of the roofline was achieved in 96% patients with an RF application time of 10 min (IQ range 8 – 13). 22 (96%) Roof dependent ATs were successfully ablated. Peritricuspid block was achieved in 96% patients with an RF application time of 12 min (IQ range 10 – 15) and all peritricuspid ATs were successfully ablated.

Among the remaining 18 patients, 10 patients had micro-reentry or focal AT involved in the LA anterior wall, 5 patients had micro-reentry or focal AT involved in the PV, 2 had AT related to LAA, and 1 had AT related the septum. Apart from 2 patients with LA anterior wall related AT, all micro-reentry or focal AT were successfully ablated.

In total, the rate of successful single mechanism AT ablation was 93% (80/86). Therefore, 64 (80%) patients with single mechanism AT required at least one of mitral line, roofline, and peritricuspid line to finally restore SR.

### Ablation of multiple mechanisms intermediate ATs

In 27 patients with multiple mechanisms ATs, 24 patients had two types of ATs and 3 had 3 types of AT. 23 patients initially presented with macro reentrant AT. 6 patients were considered having ‘dual-loop’ AT, which had both mitral and roof dependent re-entry. Among these patients, anterior LA linear lesions, which joins the mitral annulus to the right superior PV lesions, successfully terminated AT in 3 (50%) patients. The rate of conduction block across the anterior LA line was 33%. Based on the experience of the operators, all of these 6 patients received mitral line and roof line ablation.

Among 24 patients with 2 types of ATs, 17 (71%) were successfully ablated. The reason for failed ablation were unable to achieve perimitral block in 4 (57%) patients, difficult to achieve LA anterior wall block in 2 (29%), and unable to achieve peritricuspid block in 1 (14%) patient who was very fat. In 3 patients with 3 types ATs, 2 were successfully treated. One patient was initially presented as micro reentry AT in the LA anterior wall, followed by typical AFL, and finally restored to SR by superior vena cava isolation. The other patient had AT related to PV, LA roof, and finally terminated by LAA base ablation. The remaining 1 patient underwent DCCV because the anterior LA line could not be block.

Taken together, 18 (66.7%) patients with multiple mechanisms ATs restored SR by catheter ablation successfully, which is significantly lower than patients with single mechanism AT (p < 0.001). In additional, all of these 18 patients required at least one of mitral line, roofline, and peritricuspid line to finally restore SR. In total, 98 (72%) patients had successfully ablated intermediate ATs. Among these patients, 88 (90%) required at least one of the perimitral line, roofline, or peritricuspid line to finally restore sinus rhythm.

### Rhythm outcome of study population

For 136 patients who converted to organized AT during ablation, 75 (55.1%) were still in SR, 97 (71.3%) were freedom from AF recurrence, and 114 (83.8%) were freedom from AT recurrence at the end of 12 months of follow-up (Figure 2A).

**Figure 2A.**
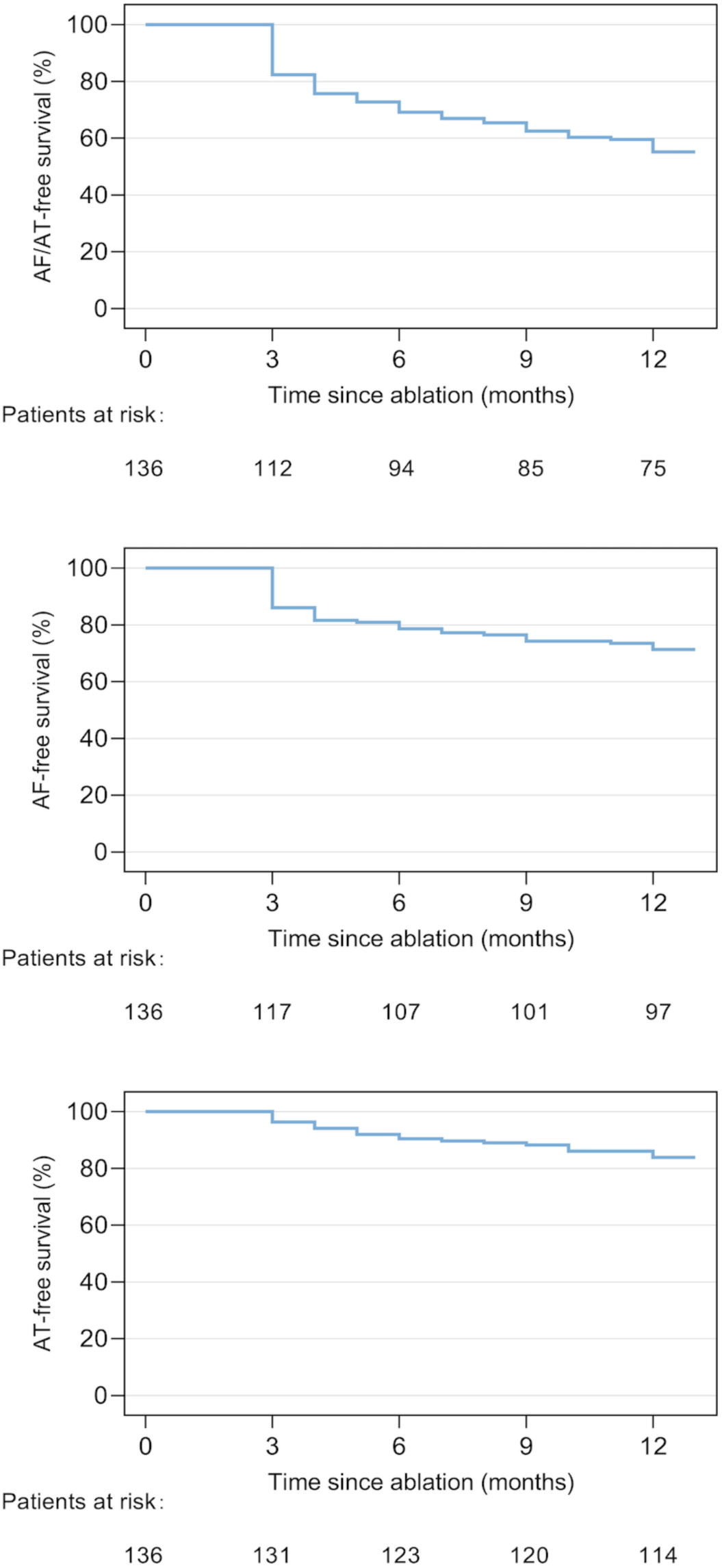
Rhythm outcome in patients converted to organized AT during ablation

As for 98 patients who finally restored SR through ablation of intermediate ATs, 63 (64.3%) patients were free of any arrhythmia, 73 (74.5%) were free of AF recurrence, and 88 (89.8%) were free from AT recurrence at the end of 12 months of follow-up (Figure 2B).

**Figure 2B.**
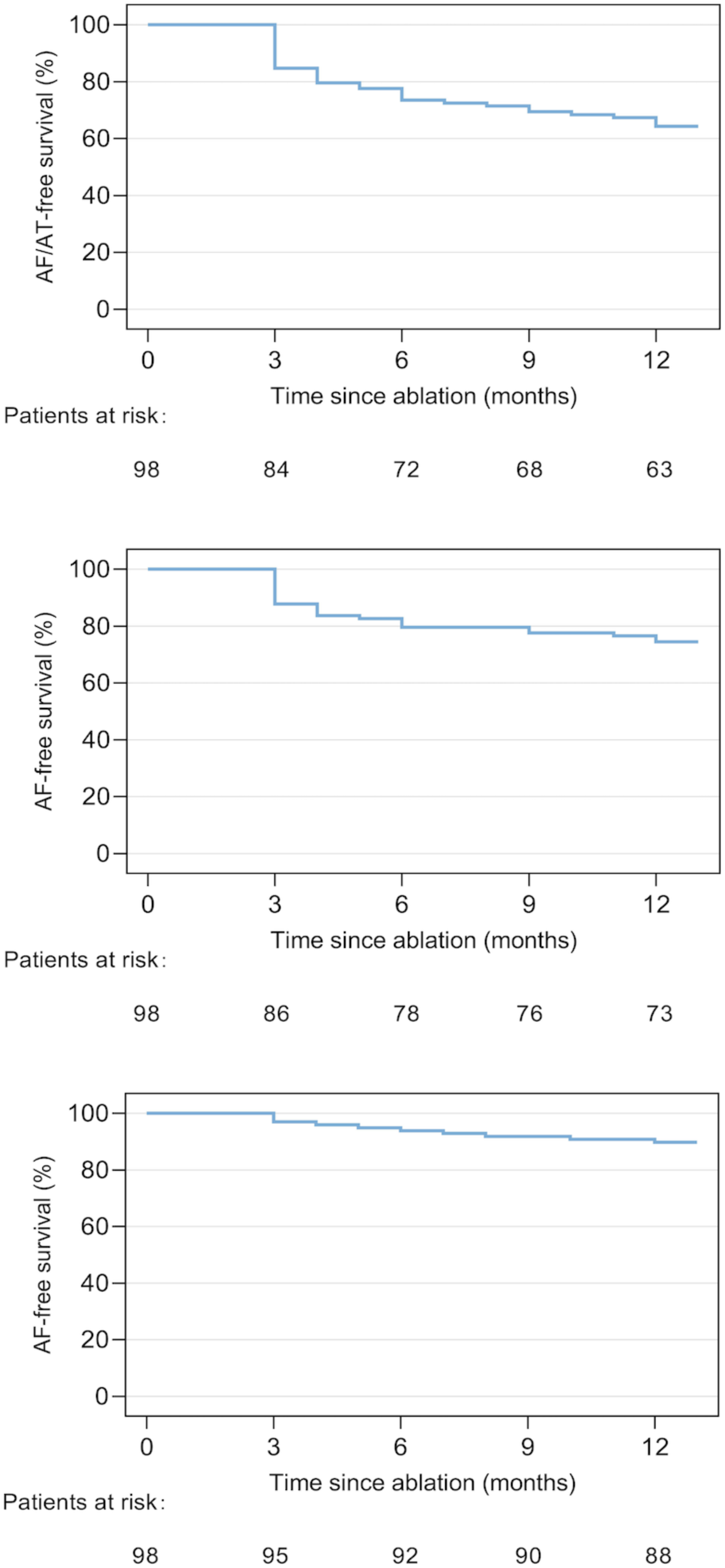
Rhythm outcome in patients restored SR by ablation of intermediate ATs

## Discussion

### Main Findings

The present study pioneered the application of the reverse thinking method, presenting empirical evidence for additional linear ablation following the mechanisms of intermediate ATs post PerAF termination. Principal outcomes are as follows: 1) ATs after AF termination can be successfully mapped (87%) and ablated (73%) in the majority of patients. and 2) Macroreentrant mechanisms representing the most prevalent in these ATs, constituting 77% of the total ATs. Of these macro-reentrant ATs, PM-AT were the most frequent, followed by RF-AT and typical AFL. Collectively, nearly 90% of patients required a minimum of one perimitral line, roofline, or peritricuspid line to successfully restore sinus rhythm, which may corroborate the efficacy of mitral line, roofline, and tricuspid line ablation.

### Linear ablation of PerAF

Our investigation that offers a unique study design employing a reverse thinking strategy to underscore the importance of linear ablation for PerAF. By comprehensively analyzing the underlying mechanisms of ATs observed during AF ablation, we strived to systematically clarify the causal relationship, thereby illuminating the rationale and imperative need for linear ablation in the management of persistent AF. Haïssaguerre et al. pioneered the introduction of linear ablation in 1994, revealing its potential by achieving successful sinus rhythm restoration in a PerAF case via the creation of linear lesions(1). Nonetheless, it warrants mentioning that despite the increasing volume of research focused on linear ablation primarily originating from the theoretical underpinnings of surgical maze procedures(9,10) that encompass atrial isolation and interruption of reentrant electrical activity, there exists a noticeable dearth of compelling evidence gleaned from rigorous, evidence-based medicine studies. Within the confines of our own study, we noted that a significant percentage (63%) of patients undergoing ablation for PerAF encountered AT during the procedure. This striking observation affirms the crucial role of linear ablation in PerAF management, offering substantial empirical evidence to corroborate its efficacy and clinical applicability.

### Mechanism of macro reentrant atrial flutter

With the escalation in catheter radiofrequency ablation procedures, the mechanism and post-ablation results of macro-reentrant AT have garnered considerable attention from electrophysiologists. Conventionally, the mechanism of macro-reentrant AT encompasses the initiation and maintenance of reentrant activation around a stable centrally located obstacle. This central obstacle can be composed of normal or abnormal structures and may be static, functional, or a combination thereof(11). In patients transitioning from AF to AT during ablation procedures or patients exhibiting post-ablation recurrence of AT, the primary mechanisms of macro-reentrant AT were PM-AT (30%-50%), RF-AT (10%-30%), and typical AFL (10%-60%) (12–15), with PM-AT ablation being the most challenging(16).

Anatomical factors, such as the presence of endocardial pouches, coronary arteries inducing a heat-sink effect, and myocardial thickness can present difficulties in achieving permanent conduction block in certain patients. Furthermore, the VOM has variable connections to neighboring myocardium, such as CS, LA, or PVs, making the achievement of isthmus block often necessitate radiofrequency energy delivery in the coronary sinus. This can be challenging due to its tortuosity and may pose a risk of injury to the coronary arteries(17). The recent VENUS study demonstrated the efficacy of additional VOM ethanol infusion. In our study, we also implemented ethanol infusion of VOM in 40% of patients with confirmed isthmus-dependent atrial flutter, resulting in a heightened isthmus block rate of 91%. Although ethanol ablation of the Marshall vein is not a universal remedy for isthmus-dependent atrial flutter, it retains an increasingly significant role.

### Mapping and ablation of other complex atrial flutter

High-density mapping has been validated by numerous prior studies as being instrumental in detecting various complex cardiac arrhythmias (18,19). In our research, high-density mapping was implemented for all patients with AT. The preliminary step during activation mapping entails the assessment the likelihood of macro-reentry. By eliminating wavefront collision around the mitral valve/roof /or tricuspid valve, PM-AT/RF-AT/or typical AFL can be respectively ruled out. Only upon the exclusion of common macro-reentrant arrhythmias, other intricate arrhythmias such as figure-of-eight reentry, double atrial reentry, and localized reentry are contemplated. In our experience, these arrhythmias typically involve critical isthmuses, resulting in profoundly slow conduction with extended duration electrograms and complex electrograms. Ablation targeting these regions usually terminates AT. Further ablation lines targeting other areas such as the mitral isthmus line and roof line may be advantageous in improving the prognosis of AT post termination of complex atrial flutter. Moreover, multiple linear lesions might precipitate the emergence of localized reentry, leading to areas with double potentials and far-field electrograms that pose consistent reconciliation challenges for automated algorithms and/or the operator. Even with a high-density map, areas of low voltage and scar in the atria harboring localized reentry might not readily yield a diagnosis. This is a commonplace scenario with small circuits and necessitates user intervention for accurate timing assignment and/or performing entrainment mapping to discern the components of the reentry circuit. The mapping and ablation of complex ATs continue to demand more precise ablation catheters and mapping systems.

### Ablation strategies for persistent atrial fibrillation

While guidelines(20) have yet to proffer conclusive recommendations regarding ablation strategies for PerAF, many electrophysiologists continue to favor linear ablation.This strategy provides a relatively fixed and straightforward ablation method aimed at achieving a “bidirectional” block and obtaining clear electrophysiological ablation endpoint, a feat challenging to achieve with other mainstream ablation strategies that lack definitive electrophysiological endpoints. However, linear ablation also presents certain limitations. Primarily, as a standardized rather than personalized method for AF ablation, it may not be suitable for all patients. Not all patients with PerAF possess reentry circuits around the mitral isthmus, tricuspid isthmus, or LA roof. Additionally, its incapacity to address the maintenance mechanism of PerAF makes it difficult to terminate AF intraoperatively(21). In our prior study, standalone linear ablation yielded an intraoperative AF termination rate of a mere 20.7%(4). Incorporating electrogram-guided ablation to linear ablation increased the termination rate to 66.7%. Significantly, patients achieving AF termination exhibited a lower incidence of AF recurrence. Therefore, extensive ablation targeting electro-anatomical mechanisms to pursue AF termination might constitute the optimal strategy for PerAF. The need for additional studies and improvements in ablation strategies remains imperative to improve outcomes in PerAF patients and alleviate the burden of this arrhythmia.

### Rhythm outcome of AT converted from AF during ablation

Patients reaching AF termination during catheter ablation are generally considered to have a superior prognosis(22,23). However, for the majority of patients with PerAF, direct conversion to SR is frequently challenging to accomplish, and an intermediate rhythm of AT is often encountered. Although initial studies implied that this arrhythmia might predict a poorer prognosis and that ablation lines are difficult to sustainably block(7), advancements in catheter ablation techniques and mapping technology have made it possible for stable and complete linear ablation of this rhythm disturbance to also lead to favorable outcomes(24). In fact, some studies(25) have shown that intervention using linear ablation targeting the mechanisms of AT have been more effective in achieving stable SR than both direct current cardioversion and direct SR, with a higher success rate sustained at the 1-year mark following the first and second procedures. In our study, among 98 patients who restored SR through ablation of intermediate AT, 63 (64.3%) patients remained free of any arrhythmia at the end of the 12-month follow-up period. This evidence suggests that favorable outcomes can be achieved by restoring SR via the ablation of intermediate rhythms.

### Study limitation

This study had several limitations. Firstly, this was a descriptive study probe into the mechanisms of intermediate ATs occurred after AF termination in the Extent-AF study. The efficacy of additional linear ablation should be further demonstrated in large randomized, control study. Secondly, in this study, we focused on intermediate ATs occurred after AF termination. Considering the various AF termination rate of different ablation strategies, studies that applied different ablation strategies may lead to different results. For example, PVI plus additional linear ablation had relative low AF termination, which was around 20% in previous literature. Thirdly, the Extent-AF study did not include a PVI alone arm, additional substrate modification increased the incidence of AT and most of them are iatrogenic ATs. It is possible that PVI alone may have some benefit over extensive substrate modification.

## Conclusions

During PerAF ablation, macro-reentrant ATs accounted for up to 77% of ATs occurred after AF termination. 90% patients required at least one linear lesion across mitral isthmus, roof and tricuspid isthmus to restore SR, which provide evidence for additional linear ablation.

## Data Availability

The data underlying this article will be shared on a reasonable request to the corresponding author.

## Acknowledgements

None.

---

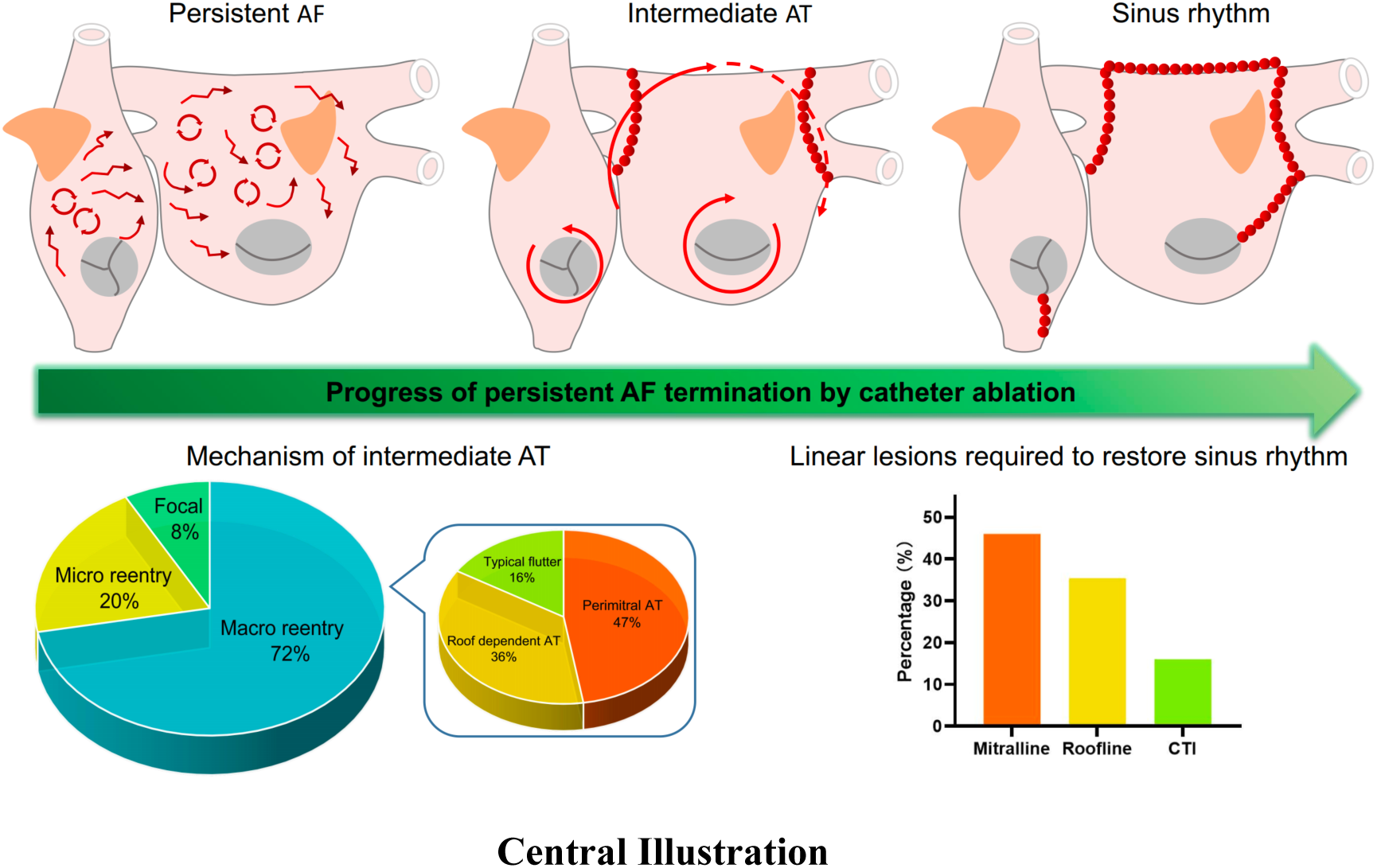

